# VAPGAMO Trial Protocol: A Cluster-Randomised Controlled Evaluation of a Digital Game-Based Learning Intervention to Reduce Adolescent Vaping Intention in Malaysian Public Secondary Schools

**DOI:** 10.1101/2025.09.04.25335146

**Authors:** Muhammad Zulhilmie Saruddin, Ahmad Zaid Fattah Azman, Rosliza Abdul Manaf, Farah Nadia Azman

**Author notes:** Correspondence to Dr Ahmad Zaid Fattah Azman.

## Abstract

**Introduction:** Youth vaping remains a global public health challenge. We will evaluate a pragmatic, theory-driven digital game-based learning intervention (VAPGAMO), delivered in public secondary schools to reduce adolescents’ intention to vape at 3-month follow-up.

**Methods and analysis:** Parallel cluster-randomised controlled trial with schools as clusters. Eight public secondary schools in an urban Southeast Asian district (Klang, Malaysia) will be randomised 1:1 to intervention or wait-list control. The intervention is a single 90-minute, facilitator-led module grounded in the Theory of Planned Behaviour. Surveys at baseline, immediately post-intervention, and 3 months. The primary outcome is intention to vape at 3 months. Secondary outcomes are knowledge, attitudes, injunctive norms, and refusal self-efficacy (perceived behavioural control). Pre-specified implementation outcomes include acceptability, fidelity, reach, time-on-task, and cost per student. Primary analysis will use generalised estimating equations with cluster-robust standard errors, adjusted for baseline covariates; intracluster correlation coefficient and design effect will be reported; intention-to-treat will be applied.

**Ethics and dissemination:** Ethical approval: Universiti Putra Malaysia JKEUPM-2024-887; approvals from the Ministry of Education, Malaysia. Findings will be disseminated in peer-reviewed outlets and shared with education and health authorities to inform school-health programming. De-identified data and code will be made available upon publication, subject to approvals.

**Trial registration:** This study was registered with the Thai Clinical Trial Registry (TCTR) [Ref No. TCTR20241222001], registered on December 22, 2024.

**Strengths and limitations of this study:**

- Pragmatic cluster-randomised design embedded in routine public schools, enhancing external validity for similar systems.
- Theory-driven digital module with pre-specified implementation outcomes to inform scale-up decisions.
- Analysis plan uses appropriate cluster methods with ICC reporting.
- Single 90-minute dose and 3-month follow-up may limit durability of effects.
- Conducted in one district; generalisability to other settings will be empirically assessed via implementation metrics.

## BACKGROUND

Adolescent vaping has emerged as a global public health threat, complicating efforts to curb nicotine addiction and tobacco-related harm among youth (1). Electronic cigarettes (ECs) or Electronic Nicotine Delivery Systems (ENDS), including cig-a-likes, pod-mods, and customizable vaporizers, are increasingly popular due to their sleek design, flavor appeal, ease of access, and discreet usage among youth worldwide (2). These devices deliver nicotine through aerosolized e-liquids, mimicking traditional smoking behaviors while exposing users to ultrafine particles, carcinogens, and other harmful chemicals (3–6). Early exposure heightens the risk of lifelong nicotine addiction and has been linked to neurodevelopmental disruption, particularly during adolescence (7–9).

The prevalence of adolescent vaping is accelerating globally. A global meta-analysis estimates that 1 in 6 adolescents have tried vaping, with 4.8% being current users (10). A nine-fold increase in daily vape use among 14–15-year-olds in New Zealand was reported between 2015 and 2023, with 10% of adolescents in this age group currently vaping (11). In the United Kingdom, experimentation with vaping is defined as trying at least once, rose by 50% from 7.7% in 2022 to 11.6% in 2023 (12). These trends underscore the urgency of addressing vaping intention among adolescents, a key predictor of initiation. Intention, or one’s cognitive motivation to engage in a behavior, is a core construct in behavior change theory and is strongly associated with future vaping uptake (13–15). For instance, a cohort study found that adolescents who intended to vape were 2.5 times more likely to initiate within six months (16).

Despite international concern, many existing school-based prevention initiatives remain focused on conventional tobacco products (17, 18). In Malaysia, adolescent vaping has surged from 1.1% in 2011 to 14.9% in 2022, with over 40% of its user initiating use before the age of 14, particularly in high-burden states like Selangor (19, 20). Current school-based programmes such as the *“Kesihatan Oral Tanpa Asap Rokok Programme” (KOTAK)* and IMFree largely emphasize traditional smoking and may not resonate with today’s digital-native adolescents (17, 18, 21). There is a growing consensus on the need for innovative, culturally relevant, and theory-driven interventions. Game-based learning (GBL), which merges interactive digital experiences with behavioral science, holds promise in shaping health behavior among adolescents. Over 90% of youth globally play video games, spending an average of 6.3 hours per week (22, 23). In Malaysia, over half of the nation’s 20.1 million gamers are school students and young adults, reflecting a unique opportunity to embed public health interventions in digital platforms. GBL has demonstrated effectiveness in influencing behaviors related to sexual health, obesity, and mental health (24–26). A recent systematic review also supports GBL’s utility in improving vaping-related knowledge, attitudes, and harm perceptions (27).

Digital interventions such as gamified learning align well with preferences of Gen Z and Gen Alpha learners, who favor fast-paced, interactive and segmented content (28). These approaches not only correct misconceptions and strengthen refusal skills, but also build intention to resist vaping (29). By providing a multi-sensory, active and experiential learning environment, gamification fosters the development of decision making and problem solving skills throughout the gameplay (30).

Moreover, embedding such interventions within school environments provides an ideal platform for scalable primary prevention (31). Grounded in the Theory of Planned Behavior and aligned with the WHO Framework Convention on Tobacco Control (FCTC), this study aims to develop and evaluate a culturally tailored, theory-based GBL intervention to reduce vaping intention among adolescents in Malaysia. This intervention targets the key cognitive antecedents of intention, which are subjective norms, attitudes, and perceived behavioral control to strengthen adolescents capacity to resist vaping initiation.

## METHODS AND ANALYSIS

### Overall study design

This is a parallel cluster-randomized controlled trial (cRCT), with schools being the unit of randomization (clusters). The trial will be used to assess the effectiveness of a game-based learning intervention program on vaping intention, knowledge, attitude, subjective norm and perceived behavior control of national secondary school students in Klang district, Malaysia. The Klang district was selected as the trial location for as it is one the largest district in Selangor and Selangor being among the top highest vape user. There are twenty-two eligible schools in Klang district. Eight of the schools were selected as clusters (schools), and randomly assigned into the intervention and wait-list control group. The intervention group will receive a GBL intervention on vaping. Those in the wait-list control group will receive the usual care first and then cross over to the intervention arm after 3-months (Figure 1).

### Eligibility Criteria

The inclusion criteria for schools will take into account the following requirements: (1) being a national secondary school; (2) they are located in Klang district; (3) and they consent to participate in the study. The exclusion criteria will disqualify national schools that are religious-based school, non-coeducational school boarding school and upper form (form four and above) or Form six only schools and school without computer lab facility from the study. The inclusion criteria for students takes into consideration the following requirements: (1) such participants must Malaysian citizen; (2) participants are aged between 13 to 15 years old; (3) they are literate in Malay or English language to take part in the study. The only exclusion criteria for students are those who are attending special education classes.

### Intervention

The intervention group will be introduced to a game-based learning intervention program on vaping intention. This intervention is grounded in the Theory of Planned Behavior (TPB) and developed by utilizing GBL approach, whereby educational elements are embedded into the interactive game-play and players would subconsciously learn while playing. Many smoking/vaping educational interventions, being grounded in the TPB, have shown improvement in avoiding vaping intention. The GBL intervention program uses four constructs of the TPB: attitude, subjective norm, perceived behavior control and behavior intention. This intervention was prepared and designed to bridge the gap of vaping knowledge and to improve vaping avoidance among these students. Table 1 gives an outline of GBL intervention on vaping intention along with the application of TPB concepts in the GBL intervention.

**Table 1.**
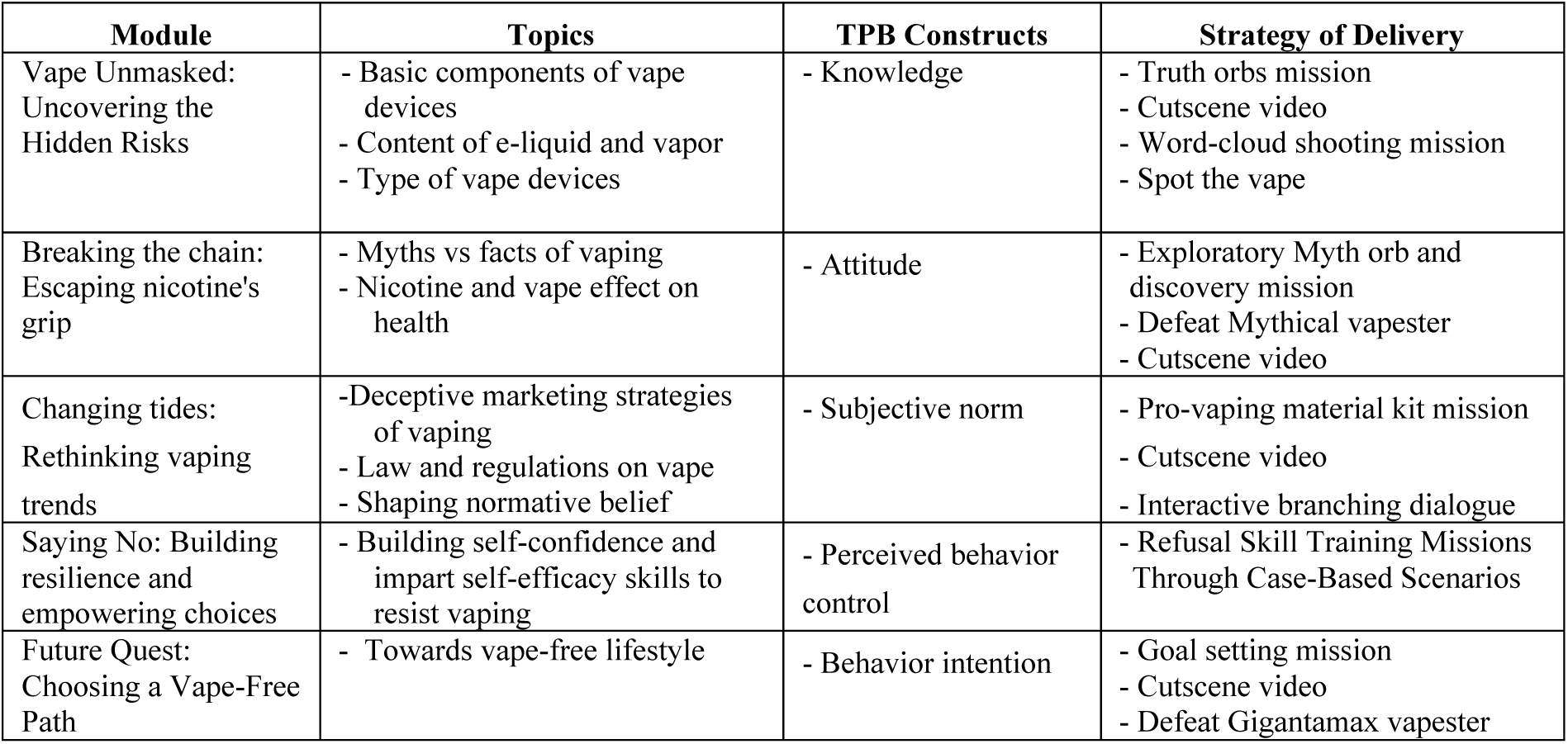
Outline of game-based learning intervention on vaping intention.

The GBL intervention consists of five modules and pending copyright protection by the Intellectual Corporation Property of Malaysia (MyIPO). Module 1 (*Vape Unmasked: Uncovering the Hidden Risks*) provides general overview of vape devices and its harmful contents in order for participants to have clear understanding of the topic. Module 2 *(Breaking the chain: Escaping nicotine’s grip)* give information on health implication of vaping, potential nicotine addiction and correct misconception about vaping. Module 3 *(Changing tides: Rethinking vaping trends)* further explore on strategy to denormalize vaping behavior, reduce social acceptability and raising awareness to discourage vaping experimentation. Module 4 *(Saying No: Building resilience and empowering choices)* will strengthen participant’s belief to resist vaping and enhance their self-efficacy to refuse usage. Module 5 *(Future Quest: Choosing a Vape-Free Path)* offers guidance to set personal goals for maintaining a vape-free lifestyle.

Following the development of the GBL intervention, face and content validity were checked and assessed by five professional expert panelists from the Community Health Department at the Faculty of Medicine and Health Sciences, Universiti Putra Malaysia, Selangor State Health Department and Klang District Health Office. The experts endorsed the educational material as effective in meeting the study’s objectives. They emphasized the importance of clarity and simplicity, recommending the use of fewer words and replacing them with more illustrations to enhance engagement and using characters that resemble the participants to help them relate better and improve understanding. They also suggested that the video game be made available in the Malay language and incorporate common teenage slang, as most young, vulnerable vapers are Malays. Additionally, they recommended incorporating scenes that depict peer influence in avoiding vape use, highlighting family role, and promoting skills that encourage good and healthy behaviors.

The GBL intervention involves a 90-minutes session for one-day. The study researcher will conduct the session for intervention group at each respective school. It is expected that four sessions will be carried out each week. Although the implementation date of the intervention may differ from one school to another, its duration time will be the same for all. Participants in the control group will not receive any education during the study period. However, they will be given the same GBL intervention materials on vaping intention prevention at the end of the study, and they also will answer the same sets of the questionnaires at baseline, immediately after the intervention, and then, three-months after the intervention. The following is a breakdown of the gameplay to be played:

a. The player assumes the role of a young teenage boy who serves as the virtual avatar, navigating the game world and progressing through challenges. Player will engage combat with the Vape-like monster (Vapester) and collect valuable items to enhance their abilities and knowledge. In a 15-minutes of level 1, player will engage combat with first and second generation vapester while collecting ‘Truth Orbs’ to gather information on vape device and its basic components. Player will encounter with three mystery boxes to unlock cut scene video on how vape device’s work and its harmful content. Other mystery boxes will unlock mini-games called ‘Word-cloud shooting mission’, that teach player about the harmful chemicals in vape and identifying various design of vape in ‘Hidden in a plain-sight’ mini game.
b. Player will progress to level 2, to combat with more difficult third and fourth generation vapester, while discovering the ‘Myth orbs’ to uncover myths and facts of vaping. Similarly, player will encounter another two mystery boxes. This will unlock a cut-scene video on nicotine and vaping effect on health. At the end of level 2, player will unlock another mini-game to test their knowledge and information gathered to defeat the 4th-generation *Mythical Vapester*.
c. In a 30-minutes level 3 game, player’s goals are to denormalize vaping and reduce its social appeal by finding and destroying the marketing kit to promote vapes. This will reveal the hidden truth behind the marketing kits and its role. Player will further uncover the truth through mystery box cut scene video on tricky marketing strategies to deceive youngster. Next mini game aim to empower player denormalize vaping through existing regulations by playing tag-match game to transforms various vape-related situation into vape-free environment. The last mini-game in this level is interactive branching dialogue. Player will engage in two settings (at home and school setting) to rethink what others are expected of them to change the tides.
d. In a final level, player will continue the exploration and collecting the goal and strategy cards to set a personal goal and aims to remain vape-free. Concurrently, player will enhance their self-efficacy and refusal skill, while refusing peers in risky social situation through series of scenario making the best choices. Lastly, player will unlock one cut-scene video to empower vape-free youth and final epic combat with ‘*Gigantamax Vapester’* by using all the information and items gather to defeat this final enemy.

### Participant Privacy

To ensure sincere responses, honesty will be emphasized to the participants. They will be informed that all questionnaires will be anonymous with a unique number to identify participants later on.

### Outcomes Measures

#### Primary Outcomes

The primary outcome variable in the present study is vaping intention. It will be measured using a modified questionnaire consisting of items adapted from Evans, Bingenheimer (32) and Noar, Rohde (33). This self-administrated questionnaire consists of four items as followed: (a) Do you think you will use vape (even one puff) within the next few months? / (b) in the next year? / (c) in the next 5 years? (d) Thinking about the future, if one of your best friends offered you a vape (even one puff) in the coming year, would you smoke it? These variables will be average of four items, which will be answered on a 5-point response scale ranged from “not at all likely” (1) to “extremely likely” (5). The reliability of the scale was good (Cronbach’s α = 0.95).

#### Secondary Outcomes

The secondary outcome variables in this study include:

##### Attitude towards vaping

Attitude towards vaping will be measured using the Electronic Cigarette Attitude Survey (ECAS) instrument. Developed by Diez, Cristello (34), this instrument incorporates 12 items pertaining to attitude for adolescent’s vape use. Participants will be asked to rate their level of agreement to these 12 statements comparing vape use to conventional cigarette. Each item will be coded on a Likert-scale ranging from one to five with 1= *“strongly agree“* and 5= *“strongly disagree”.* The average mean score will be calculated and higher score indicates positive attitude. The internal consistency for the overall measure was good (Cronbach’s α = 0.93).

##### Subjective norm of vaping

Subjective norm of vaping will be assessed by questionnaire consisting of items adapted from Lei, Qi (35). The questionnaire consists of three items as followed: (a) Most people who are important to me think I should use vapes to instead of conventional cigarettes; (b) Most people who are important to me would want me to use vape; (c) People whose opinions I value would prefer that I use vape. Each item will be rated through five-point Likert scale ranging from strongly disagree (1) to strongly agree (5). All items have good internal consistency (Cronbach’s alpha: 0.96).

##### Perceived behaviour control

Perceived behavior control from vaping behavior will be assessed by items adapted from Lei, Qi (35). The questionnaire also consists of three items as followed: (a) Whether or not I use vape is entirely up to me; (b) I am confident that if I want, I can buy and use vape; (c) I have resources, time and opportunities to buy and use vape. Each item will be rated through five-point Likert scale ranging from strongly disagree (1) to strongly agree (5). All items have good internal consistency (Cronbach’s alpha: 0.94).

For interpretability, subjective norm and perceived behavioural control items will be reverse-coded so that higher scores indicate greater social disapproval of vaping and higher refusal self-efficacy.

##### Knowledge of vaping

Knowledge of vaping will be measured using a modified questionnaire consisting of items adapted fromRohde, Noar (36), Hafiz, Rahman (37), Le, Tran (38). Participants are required to determine whether each statement is “true,” “false,” or “don’t know.” Total of 20 items will be used to assess knowledge of vaping. Each correct answer received “one point” and incorrect/ “Don’t know” response received “zero points”. The scale score ranged from 0 to 20. The level for knowledge was measured by the sum of the score obtained, with a higher score indicating greater knowledge.

#### Other outcomes

##### Participants’ Personal Information

This part of the questionnaire consists of five questions conducive for assessing the sociodemographic factors of the participants (i.e., age, gender, ethnicity, academic form, school location).

Lawshe’s method was used to examined the content validity index (CVI) of the questionnaire, which aimed to assess the clarity, relevance, and cultural appropriateness of each construct using the item-level content validity index (I-CVI). This self-administered questionnaire was reviewed by five professional expert panelists, comprising three public health physicians from the Community Health Department at the Faculty of Medicine and Health Sciences, Universiti Putra Malaysia, one family medicine specialist from Ministry of Health (MOH), and one secondary school teacher from Ministry of Education (MOE). According to Lynn (39), an acceptable I-CVI should not be lower than 0.78. In addition, the scale-level content validity index (S-CVI) was calculated to determine the proportion of items deemed content valid across the entire scale. Waltz, Strickland (40) recommended an S-CVI value of 0.90 or higher. The I-CVI scores for all constructs ranged from 0.93 to 1.00, while the S-CVI scores ranged from 0.92 to 1.00. The face validity of the questionnaire was initially done by the ten target participants, who were not included in the study. Then some changes on the relevant items were made depending on the suggestions provided by these professional experts and every students feedback.

#### Implementation outcomes

We will assess five implementation outcomes: acceptability (post-session 5-item student rating, 5-point Likert, higher=more acceptable), fidelity (facilitator checklist of core components delivered; % of items delivered), reach (proportion of eligible students who attend the session), engagement (time-on-task and levels completed from in-game logs), and cost per student (micro-costing of personnel time, materials, and overhead). These will be summarised descriptively with 95% CIs and compared between arms as exploratory analyses.

### Translation of the Questionnaire and Study Module

The questionnaire and study module were translated according to the process of professional translation, while the back translation was used as an approach for the adaptation of instruments. These instruments were originally administered in English. The translation process into the target Malay language for use in this study will follow the steps outlined below.

i. Forward translation to Malay language by two native speaker of Malay language and fluent in English language, any discrepancy will be discussed and resolved
ii. Back-translation to English language by another two independent translators who are proficient in both English and Malay language and had no prior knowledge on the questionnaire and module
iii. The back-translation will be compared with original questionnaire and module to ensure accuracy of the forward translation.
iv. Pre-testing of the translated questionnaire and module among 30 secondary school students, who are not of the participants in the study. The clarity, understandability, and quality of the module and questionnaire were discussed with them individually. Then, phrases that were unsuitable and difficult to understand were identified, and accordingly, adjustments and modifications thereto were made following their evaluation.
v. Subsequently, the final version was approved for the study.

### Patient and public involvement

Patients and/or the public were not involved in the design, or conduct, or reporting, or dissemination plans of this research.

### Sample Size

For the individually randomised design, the required sample was first estimated using a two-means comparison (power 0.80, two-sided α=0.05) with μ₁=4.26 and μ₂=3.91 (41). We then inflated by the cluster design effect, DE=1+(m−1) × ICC. Assuming an average cluster size m≈46 across eight schools and ICC=0.009, DE≈1.4. Allowing 10% attrition yields n=366 participants across 8 clusters. We will report the observed ICC and design effect in trial results.

### Sampling Method

A total number of eight schools with 366 participants that have met the inclusion and exclusion criteria will take part in the study. Following cluster sampling, four of the schools will be involved in intervention group, while the other four are the control group. Then the proportionate allocation number of students to participate in the study will be calculated based on the density of the students in each school (probability proportional to size (PPS). Participants selection will be conducted by using a simple random sampling technique.

### Participant Recruitment

Forty national secondary schools were assessed for eligibility. There were 18 schools were not eligible to take part in the study as they did not meet the selection criteria for school by being religious-based school, non-coeducational school, boarding school, consist of upper form or Form six only school and school without computer lab facility.

With respect to the 22 eligible schools, eight schools were randomly selected and adequate to contribute the necessary sample size for this study. Written permission will be obtained from respective education authorities in the ministry, state and district level. With the help of school administrators, the school coordinator, counsellor and information and technology (ICT) teacher would be invited to attend small meeting groups in each school. During such meetings, the researcher will explain the objectives and advantages of the study, along with the inclusion and exclusion criteria. After selecting the required participants’ number in each school, participants will be asked for written informed consent. Accordingly, participants who agree to participate will be asked to fill in the questionnaire. Later on, educational interventions (GBL) on vaping intention will be delivered to the intervention group. Following that, the participants will be requested to fill in the same set of the questionnaires (with the exception of the personal information form) in the post-immediate and follow-up assessment.

### Study Principles

The reporting of this protocol followed the Standard Protocol Items: Recommendations for Interventional Trials (SPIRIT) 2013 Statement (41). The study reporting will be in accordance with the Consolidated Standards of Reporting Trials (CONSORT) statement. Figure 2 below illustrates the Consolidated Standards of Reporting Trials (CONSORT) flowchart (42).

**Figure 2.**
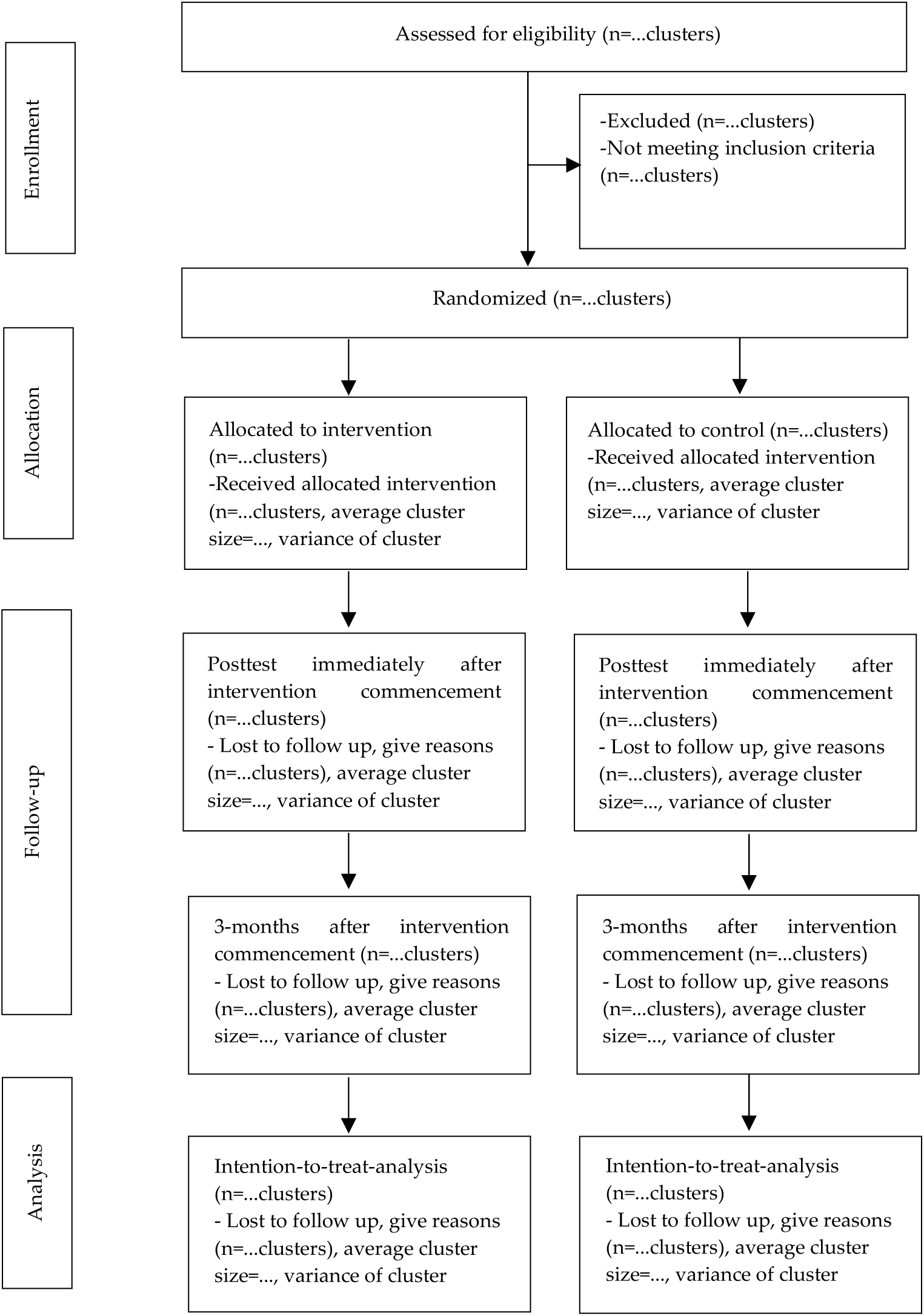
CONSORT flow diagram of the study—adapted from Campbell et al. (42)

### Randomization

The unit of randomization in this current study is a school. To allocate the eight selected clusters (schools) to the study groups, each school will be assigned a number one to eight. Simple randomization technique will be done using online random group allocation to create the randomization sequence, with 1:1 allocation. Thus, four clusters were randomly allocated to the intervention group (SR3, JK5, CV2, and RP7) and four clusters to the control group (TJ8, SA6, MK4, and TG1).

### Allocation Concealment Mechanism

To ensure proper allocation concealment, an independent assistant will be assigned to produce the allocation sequence list. Each cluster will be given a unique code in a sealed opaque envelope. Following that, the assistant will open the envelopes and assign the clusters to the intervention or control group based on the list of codes generated by the software.

### Blinding

Single blinding will be applied in this study. The group that should receive the health intervention should be made known to researchers due to their involvement in study implementation. The participants will be blinded for the study to keep participant unaware of their group status to minimize performance bias.

### Statistical Analyses

Data will be analyzed using Statistical Package for the Social Sciences (IBM SPSS, Version 29.0). We will analyse data in SPSS under an intention-to-treat framework. The primary outcome (intention to vape at 3 months) will be modelled using generalised estimating equations with an exchangeable working correlation and cluster-robust standard errors, adjusting for baseline outcome and prespecified covariates (age, sex, school). Continuous secondary outcomes will use the same approach. We will report adjusted mean differences with 95% CIs, the intracluster correlation coefficient, and the design effect. Missing outcome data will be handled using multiple imputation if >5%; a complete-case analysis will be a sensitivity check. A per-protocol sensitivity analysis will be conducted based on session completion.

## Discussion

This study presents a protocol for evaluating a theory-driven game-based learning intervention program (GBLIP) aimed at reducing vaping intention among national secondary school students in Selangor, Malaysia. Given that a significant proportion of adolescent vapers report intention to vape before the age of 14, the findings underscore the urgency of addressing early susceptibility to nicotine use in line with broader tobacco control efforts. By integrating digital technology, behavioral theory, and school-based delivery, this intervention offers a promising pathway to reshape health communication for digital-native adolescents.

Crucially, the GBLIP model (VAPGAMO) aligns with international tobacco control policy frameworks, particularly the WHO Framework Convention on Tobacco Control (FCTC). Specifically, this intervention operationalizes elements of Article 12, which calls for education, communication, training, and public awareness, and supports Article 14 by contributing to demand-reduction strategies targeting tobacco and nicotine dependence. The use of interactive digital content, which is designed to improve knowledge, shift social norms, and enhance behavioural control, embodies a modernized approach to achieving these global objectives.

The significant improvements expected in knowledge, attitudes, subjective norms, and perceived behavioural control can inform national-level curriculum reform and adolescent-focused public health campaigns. Moreover, the intervention’s culturally tailored yet modular design makes it scalable and adaptable to diverse international contexts, particularly in low- and middle-income countries facing rising adolescent vape use. Although this study was conducted in Malaysia, the GBLIP model is not country-specific; its underlying structure anchored in behavioural science and gamified delivery, which can be adopted or localized by ministries of health, school health units, or international NGOs in settings with similar youth risk profiles.

Beyond demonstrating statistical significance, the study aims to generate actionable evidence for education ministries, school administrators, and youth-focused public health organizations. Policymakers may consider integrating such digital tools into existing tobacco control programs to expand their reach and impact among adolescents. Health professionals and school stakeholders can use the content to spark discussions, reinforce refusal skills, and counter the normalization of vaping culture in social media and peer groups. Limitations include a single 90-minute dose, a 3-month follow-up, and self-reported outcomes, with potential inter-school contamination. We will mitigate these through cluster-appropriate analysis, fidelity checks, and implementation metrics (reach, engagement, cost).

This study is also among the first in Southeast Asia, to the best of the researchers’ knowledge, to evaluate a video game–based intervention grounded in the TPB targeting vaping intention among adolescents. While long-term evidence on gamified interventions in tobacco control remains limited, the early implementation of such innovative approaches is critical to preempt the vaping epidemic among future generations. By promoting not only knowledge acquisition but also critical thinking, digital resilience, and decision-making skills, GBLIP fosters more than just behavioural intent as it nurtures personal agency and informed resistance in the face of aggressive nicotine marketing tactics.

In summary, this GBLIP study contributes to the global evidence base on digital strategies for tobacco demand reduction. If proven effective, it offers a cost-effective, scalable, and engaging educational tool that addresses the complex interplay of beliefs, social influences, and behavioural control in adolescent vaping. Such innovations are essential for accelerating global progress toward WHO FCTC goals, especially in reaching hard-to-engage youth populations.

## Data Availability

Data will be made available upon request and subject to an established data sharing agreement.

## Ethics and dissemination

This trial protocol has been approved by the Ethics Committee of Universiti Putra Malaysia *(JKEUPM)* [Ref No. JKEUPM-2024-887]. Written informed consent will be obtained from participants prior to enrolment. We will communicate with investigators, trial participants, trial registries and journals and submit related reports following the rules of the ethics committee for any important protocol modification during the trial process. All procedures in this study will be carried out in accordance with the ethics committee guidelines and regulations and the study protocol will be conducted in accordance with the Declaration of Helsinki and the Malaysian Good Clinical Practice Guideline. Results and findings of this trial will be disseminated in peer-reviewed journals and professional conferences.

## Ethics statements

### Patient consent for publication

This protocol paper does not contain any information about specific individuals. The study described in the paper will collect information about individuals, but any resulting published work will not identify them.

## Availability of Data and Materials

This protocol paper does not include any data analysis. Dr. Rosliza will have full access to all data collected throughout the cluster randomized controlled trial and will be responsible for ensuring the integrity and accuracy of the data and its analysis. Data will be made available upon request and subject to an established data sharing agreement.

## Authors’ Contributions

MZS and AZFA is responsible for the provision of the literature review, study design, participant’s recruitment, data collection, data analysis, and manuscript preparation. AZFA and RAM coordinated the study design and manuscript preparation. While RAM and FNA reviewed the content of the intervention material and ethical issue, AZFA planned the statistical analysis. RAM, AZFA and FNA contributed to the protocol design and reviewed and edited the final manuscript. All authors have read and agreed to the published version of the manuscript.

## Disclaimer

This funding source was not involved in study design, collection, analysis and interpretation of data, writing the manuscript and the decision to submit the paper for publication.

## Acknowledgements

The authors wish to acknowledge the Selangor State Education Department and Klang District Education Office for their dedicated help and assistance. We also thank the schools that took part in this study.

## References

1. Birdsey J, Cornelius M, Jamal A, Park-Lee E, Cooper MR, Wang J, et al. Tobacco Product Use Among U.S. Middle and High School Students — National Youth Tobacco Survey, 2023. MMWR Morb Mortal Wkly Rep 2023. 2023(72):1173–82.

2. Yang Q, Liu J, Lochbuehler K, Hornik R. Does Seeking e-Cigarette Information Lead to Vaping? Evidence from a National Longitudinal Survey of Youth and Young Adults. Health Communication. 2019;34(3):298–305.

3. WHO. Electronic nicotine and non-nicotine delivery systems: a brief. Copenhagen: World Health Organization. Regional Office for Europe; 2020. Contract No.: WHO/EURO:2020-4572-44335-62638.

4. Yimsaard P, McNeill A, Yong H-H, Cummings KM, Chung-Hall J, Hawkins SS, et al. Gender Differences in Reasons for Using Electronic Cigarettes and Product Characteristics: Findings From the 2018 ITC Four Country Smoking and Vaping Survey. Nicotine & Tobacco Research. 2020;23(4):678–86.

5. Lindpere V, Winickoff JP, Khan AS, Dong J, Michaud TL, Liu J, et al. Reasons for E-cigarette Use, Vaping Patterns, and Cessation Behaviors Among US Adolescents. Nicotine & Tobacco Research. 2022;25(5):975–82.

6. Walley SC, Wilson KM, Winickoff JP, Groner J. A Public Health Crisis: Electronic Cigarettes, Vape, and JUUL. Pediatrics. 2019;143(6).

7. Chan GCK, Stjepanović D, Lim C, Sun T, Shanmuga Anandan A, Connor JP, et al. Gateway or common liability? A systematic review and meta-analysis of studies of adolescent e-cigarette use and future smoking initiation. Addiction. 2021;116(4):743–56.

8. Fadus MC, Smith TT, Squeglia LM. The rise of e-cigarettes, pod mod devices, and JUUL among youth: Factors influencing use, health implications, and downstream effects. Drug and Alcohol Dependence. 2019;201:85–93.

9. Leslie FM. Unique, long-term effects of nicotine on adolescent brain. Pharmacology Biochemistry and Behavior. 2020;197:173010.

10. Salari N, Rahimi S, Darvishi N, Abdolmaleki A, Mohammadi M. The global prevalence of E-cigarettes in youth: A comprehensive systematic review and meta-analysis. Public Health in Practice. 2024;7:100506.

11. Egger S, David M, McCool J, Hardie L, Weber MF, Luo Q, et al. Trends in smoking prevalence among 14–15-year-old adolescents before and after the emergence of vaping in New Zealand; an interrupted time series analysis of repeated cross-sectional data, 1999–2023. The Lancet Regional Health – Western Pacific. 2025;56.

12. ASH. Use of e-cigarettes among young people in Great Britain: Action on Smoking and Health; 2023. Accessed February 1, 2024. [Available from: https://ash.org.uk/resources/view/use-of-e-cigarettes-among-young-people-in-great-britain.

13. Ulker-Demirel E, Ciftci G. A systematic literature review of the theory of planned behavior in tourism, leisure and hospitality management research. Journal of Hospitality and Tourism Management. 2020;43:209–19.

14. Scheinfeld E, Crook B, Perry CL. Understanding Young Adults’ E-cigarette Use through the Theory of Planned Behavior. Health Behav Policy Rev. 2019;6(2):115–27.

15. Nicksic NE, Barnes AJ. Is susceptibility to E-cigarettes among youth associated with tobacco and other substance use behaviors one year later? Results from the PATH study. Prev Med. 2019;121:109–14.

16. Gaddy MY, Vasquez D, Brown LD. Predictors of e-cigarette initiation and use among middle school youth in a low-income predominantly Hispanic community. Frontiers in Public Health. 2022;10.

17. Abdul Halim NA, Wee LH, Mohd Saat NZ, Jit Singh SJ, Siau CS, Chan CMH. Application of the Logic Model to the School-Based Fit and Smart Adolescent Smoking Cessation Programme. Malays J Med Sci. 2022;29(5):133–45.

18. MOH. National Strategic Plan for Tobacco Control 2015-2020. 2015.

19. NHMS. Non-Communicable Disease: Risk Factors and Other Health Problems. 2019. Report No.: ISBN: e978-967-18159-2-2.

20. NHMS. The National Health and Morbidity Survey (NHMS): Adolescent Health Survey 2022. 2022. Report No.: MOH/S/IKU/199.23(RH)-e.

21. Bernama. Education ministry to expand anti-smoking programme at primary school level. Malaysia Now. 2023 28 April 2023;Sect. News.

22. Anderson M, Jiang, J. Teens’ Social Media Habits and Experiences. Pew Research Center. 2018. Accessed November 12, 2024. [Available from: https://www.pewresearch.org/internet/2018/05/31/teens-social-media-technology-2018/

23. Alrahili N, Alreefi M, Alkhonain IM, Aldakhilallah M, Alothaim J, Alzahrani A, et al. The Prevalence of Video Game Addiction and Its Relation to Anxiety, Depression, and Attention Deficit Hyperactivity Disorder (ADHD) in Children and Adolescents in Saudi Arabia: A Cross-Sectional Study. Cureus. 2023;15(8):e42957.

24. Martin P CL, Gottot S, Bourmaud A, de La Rochebrochard E, Alberti C. Participatory Interventions for Sexual Health Promotion for Adolescents and Young Adults on the Internet: Systematic Review. J Med Internet Res 2020. 2020;22(7).

25. Barnes S, Prescott J. Empirical Evidence for the Outcomes of Therapeutic Video Games for Adolescents With Anxiety Disorders: Systematic Review. JMIR Serious Games. 2018;6(1):e3.

26. Ameryoun A, Sanaeinasab H, Saffari M, Koenig H. Impact of Game-Based Health Promotion Programs on Body Mass Index in Overweight/Obese Children and Adolescents: A Systematic Review and Meta-Analysis of Randomized Controlled Trials. Childhood Obesity. 2018;14(2):67–80.

27. Mylocopos G, Wennberg E, Reiter A, Hébert-Losier A, Filion KB, Windle SB, et al. Interventions for Preventing E-Cigarette Use Among Children and Youth: A Systematic Review. American Journal of Preventive Medicine. 2023.

28. Ramirez KP. A Systematic Review of Gen Z’s Learning Characteristics And Preferences. ResearchGate; 2018.

29. Weser V, Duncan L, Pendergrass T, Fernandes C, Fiellin L, Hieftje K. A quasi-experimental test of a virtual reality game prototype for adolescent E-Cigarette prevention. Addictive Behaviors. 2021;112:106639.

30. Adachi PJC, Willoughby T. More Than Just Fun and Games: The Longitudinal Relationships Between Strategic Video Games, Self-Reported Problem Solving Skills, and Academic Grades. Journal of Youth and Adolescence. 2013;42(7):1041–52.

31. Lenk KM, Erickson DJ, Forster JL. Trajectories of Cigarette Smoking From Teens to Young Adulthood: 2000 to 2013. American Journal of Health Promotion. 2018;32(5):1214–20.

32. Evans WD, Bingenheimer J, Cantrell J, Kreslake J, Tulsiani S, Ichimiya M, et al. Effects of a Social Media Intervention on Vaping Intentions: Randomized Dose-Response Experiment. J Med Internet Res. 2024;26:e50741.

33. Noar SM, Rohde JA, Prentice-Dunn H, Kresovich A, Hall MG, Brewer NT. Evaluating the actual and perceived effectiveness of E-cigarette prevention advertisements among adolescents. Addictive Behaviors. 2020;109:106473.

34. Diez SL, Cristello JV, Dillon FR, De La Rosa M, Trucco EM. Validation of the electronic cigarette attitudes survey (ECAS) for youth. Addict Behav. 2019;91:216–21.

35. Lei W, Qi Z, Meng-Ru C, Philip Pong Weng W. Use and Perceptions of Electronic Cigarettes among Young Chinese Generation: Expanding the Theory of Planned Behaviour. International Journal of Humanities, Management and Social Science (IJ-HuMaSS). 2022;5(1).

36. Rohde JA, Noar SM, Horvitz C, Lazard AJ, Cornacchione Ross J, Sutfin EL. The Role of Knowledge and Risk Beliefs in Adolescent E-Cigarette Use: A Pilot Study. International Journal of Environmental Research and Public Health. 2018;15(4):830.

37. Hafiz A, Rahman MM, Jantan Z. Factors associated with knowledge, attitude and practice of e-cigarette among adult population in KOSPEN areas of Kuching district, Sarawak, Malaysia. 2019.

38. Le HTT, Tran ATV, Nguyen AQ, Tran TTT. E-Cigarette Use among University Students from One University in Hanoi, Vietnam, and Associated Factors. Asian Pac J Cancer Prev. 2022;23(11):3649–55.

39. Lynn MR. Determination and Quantification Of Content Validity. Nursing Research. 1986;35(6).

40. Waltz CF, Strickland OL, Lenz ER, Satyshur RD, Stone KS, Frazier SK, et al. Measurement in Nursing and Health Research. 5 ed. New York: Springer Publishing Company; 2017.

41. Chan AW, Tetzlaff JM, Gøtzsche PC, Altman DG, Mann H, Berlin JA, et al. SPIRIT 2013 explanation and elaboration: guidance for protocols of clinical trials. Bmj. 2013;346:e7586.

42. Campbell MK, Piaggio G, Elbourne DR, Altman DG. Consort 2010 statement: extension to cluster randomised trials. BMJ : British Medical Journal. 2012;345:e5661.

